# Trajectories of COVID-19: a longitudinal analysis of many nations and subnational regions

**DOI:** 10.1101/2023.01.19.23284768

**Authors:** David Burg, Jesse H. Ausubel

## Abstract

The COVID-19 pandemic is the first to be rapidly and sequentially measured by nation-wide PCR community testing for the presence of the viral RNA at a global scale. We take advantage of the novel “natural experiment” where diverse nations and major subnational regions implemented various policies including social distancing and vaccination at different times with different levels of stringency and adherence. Initially, case numbers expanding exponentially with doubling times of ∼1-2 weeks. In the nations where lockdowns were not implemented or ineffectual, case numbers increased exponentially but then stabilized around 10^2^-to-10^3^ new infections (per km^2^ built-up area per day). Dynamics under strict lockdowns were perturbed and infections decayed to low levels. They rebounded following the lifting of the policies but converged on an equilibrium setpoint. Here we deploy a mathematical model which captures this behavior, incorporates a direct measure of lockdown efficacies, and allows derivation of a maximal estimate for the basic reproductive number *R*_o_ (mean 1.6-1.8). We were able to test this approach by comparing the approximated “herd immunity” to the vaccination coverage observed that corresponded to rapid declines in community infections during 2021. The estimates reported here agree with the observed phenomena. Moreover, the decay rates *d* (0.4-0.5) and rebound rates *r*_0_ (0.2- 0.3) were similar throughout the pandemic and among all the nations and regions studied. Finally, a longitudinal analysis comparing multiple national and regional results provides insights on the underlying epidemiology of SARS-CoV-2 and lockdown and vaccine efficacy, as well as evidence for the existence of an endemic steady state of COVID-19.

## Introduction

Quantitative studies of viral infection in human severe acute respiratory syndrome coronavirus 2 (SARS-CoV-2) infected subjects have been enabled by the massive global deployment of sensitive and rapid PCR testing for detecting viral RNA in infected persons. Data obtained with these procedures have allowed for extensive mathematical modeling of infection expansion dynamics [1]. Indeed, epidemiological modeling of this pandemic has exploded, though results have been mixed and proved how difficult it can be to provide accurate information and predictions, especially in the early stages of the pandemic [2].

COVID-19 cases initially grew exponentially in every nation. Reduction of community infection was initially achieved by non-pharmaceutical and social distancing interventions [3,4]. The drastic social distancing measures undoubtedly curbed viral expansion [5]. However, the underlying biological, environmental and social dynamics were not fundamentally modified, and viral circulation was only temporarily inhibited. National vaccination programs deployed during 2021 were also aimed to block person-to-person infection. These interventions were enacted at different times, with different levels of enforcement, compliance and extent among nations and in major regions within nations. This global “natural experiment” makes the COVID-19 pandemic a unique opportunity to longitudinally model epidemiological dynamics.

COVID-19 modeling is primarily based on the standard SIR model as the foundational tool of mathematical epidemiology and attempts to capture the main characteristics of the complex interplay among the virus, its host and the environment [6]. The theoretical SIR model’s solution converges on a logistic-like s-curve trajectory with rapid expansion reaching a peak and declining in one wave [7]. Many much more elaborate models were deployed to study COVID-19 dynamics [8,9]; however, complexity invokes problems such as overfitting, global optimization, and interpretability. An important feature not reproduced in these models is the existence of a non-trivial equilibrium setpoint.

A key criterion of epidemic expansion is the basic reproductive number (*R*_o_) which represents a disease’s transmissibility. Specifically, it is the average number of productive secondary infections arising from one active infectious individual [10]. It is derived from the ratio between the infection and removal rate constants in the SIR or similar models [11]. A bifurcation threshold condition for the occurrence of a sustained epidemic is *R*_o_≥1, meaning that as *R*_o_<1 the infection will converge on the disease-free state. This is also an indication for “herd immunity” [12,13]. In contrast to the outcome of a disease-free state, most models in the context of COVID-19 have lacked capacity to depict sustained endemic levels of infection.

Estimation of the value of *R*_o_ is commonly based on the initial exponential growth rate [14] and the median infectious period [15,16]. This is clearly an overestimate as it disregards the removal rate of cases [17]. Another problem is it ignores the distinctive infection peak and inherent inevitable negative second derivative predicted by SIR models. Other approximations treat reproductive rates as a function of time during the epidemic. Wallinga & Lipsitch [18] summarize the main methods to calculate this time-dependent “effective” *R* (*R*_*e*_). A recent review demonstrated that Cori et al. [19] derived an accurate estimate for this parameter [20]. It has also been suggested that a simple Dirac delta distribution can be used as a proxy for *R*_*e*_ [21]. These are important though *R*_*e*_ will fluctuate as a function of the changes in infection rates as the epidemic develops [22], but further discussion is beyond the scope of this paper. While these measure changes in infection rates change over time (e.g., the first derivative) they do not capture the underlying fundamental biological and social interactions.

This paper highlights applicability of mathematical models based on the viral dynamics paradigm [23–25]. A notable characteristic of these models is a non-trivial non-zero infection dynamical steady state equilibrium setpoint. Further, they represent effects of interventions to block transmission of the pathogen throughout the population. The major advantage of this methodology is the ability to derive estimations for the values of model parameters directly from the data [26].

We refrain from exploring the dynamics of the COVID-19 virus itself. SARS-CoV-2, the virus that causes COVID-19, keeps changing and accumulating mutations in its genetic code. Some variants emerge and disappear, while others emerge, spread, and replace previous variants. For the USA, for example, variant proportions are tracked at https://covid.cdc.gov/covid-data-tracker/#variant-proportions. Obviously, the strategies for suppression can interact with the evolution of the virus. We simply assume a virus able to evolve so that it can reinfect previously infected individuals.

Publicly available data for COVID-19 were used to characterize the epidemiological dynamics of community infection. The implementation of efficacious social distancing and lockdown interventions instituted across many nations allows the modeling of the dynamics of infection decay and subsequent rebound as interventions were lifted or lose effectiveness. A longitudinal comparison among nations and major subnational regions provides insights into pathogenesis that would be difficult or impossible to obtain in past pandemics.

## Materials and Methods

### Epidemiological data

Data for confirmed active infected cases, COVID-19-associated mortality and PCR tests were retrieved [27]. For most purposes we stop in September 2021 when the widespread availability of self-testing reduces the reliability of some of the relevant time series. Preliminary review shows that the data exhibit two artifacts. First, a weekly cycle is clearly observed with a tendency for more reporting in the middle of the week and less during weekends, sometimes with orders-of-magnitude differences. Second, large inter-day fluctuations are reported, sometimes with differences spanning multiple orders-of-magnitude. While it is common to smooth the data with a moving average, the resulting estimates are highly sensitive to the fitting window, especially with small numbers and the extremely noisy data (up to an order-of-magnitude between days). Therefore, weekly averages were adopted here and calculated from the geometric mean of the daily measurements to stabilize the variance in the data [28].

There is clearly a delay between time of infection and reporting. Incubation times for COVID-19 are 6.2 days and the mean generation interval is 6.7 days, with a concurrent latent period of 3.3 days [29]. Further, there is a lag between infection and detection by lab test with a skewed distribution [30,31]. While the exact value is unknown, it will only offset the data in time and does not affect the shape of the infection trajectories. Therefore, a ten-day delay is applied here to all confirmed case numbers, only shifting them left in time and not affecting the shape of the data.

#### Inclusion criteria

Analyses were performed for nations and major subnational regions with 10-fold mean difference between PCR tests and positive confirmed cases, high GDP (PPP) per capita [32] indicating the ability to perform an extensive testing program, and approximately one log decrease in infections from peak to minimum rates during lockdowns. The forty-five units qualifying are 24 European nations, Australia and New Zealand, the UK and the four nations constituting the UK, 10 USA states, and four Asian nations.

### Lockdown interventions, mobility and vaccination coverage

Dates for national policy lockdown initiation and termination are available and collated from numerous sources and the COVID-19 stringency index was accessed [33]. Even so, compliance was imperfect, and mobility was used as a minimal estimate for the efficacy of the intervention to block infection [34,35]. The number of doses of vaccines were retrieved from Mathei et al. [36] and population data from the World Bank [37]; these enable calculation of the percent of the populace vaccinated. To compare countries and regions, data are commonly normalized to population size, such as “per million.” However, COVID-19 is strongly dependent on population density [38]. Therefore, to alleviate the population density bias, the data were normalized to the built-up area [39,40].

### Mathematical modeling of COVID-19

The epidemiology of COVID-19 was analyzed using a mathematical model of viral dynamics. The three model compartments include susceptibles (*S*), COVID-19-confirmed individuals (*I*), and free virus particles (*V*). The model assumes that uninfected people are being made available at a constant rate (*σ*) and the virus productively infects them with probability *βVS*. Detected individuals are removed by quarantine at rate *δI*. Deaths can be thought of as a subset of these and are neglected for the purposes of this study. Viral particles are released from infected individuals at rate *pI* and are inactivated at rate *cV*. These assumptions lead to the coupled nonlinear ordinary differential equations:

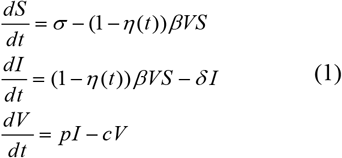

Intervention efficacy to block infection, via lockdowns or vaccination, is parameterized here by *η*(*t*). Assuming partial and incomplete effectiveness, e.g. 0<*η*<1, the system will converge on a new lower steady state. The mean infectious time is 1/*δ*. The average number of virus particles produced during the infectious interval of a single infected person (the burst size) is given by *p*/*c*. While asymptomatic carriers are thought to be efficient spreaders, they are neglected here, and we assume as a first approximation that their dynamics are similar with *I* and change in tandem with the confirmed cases.

Sustained viral propagation ensues if, and only if, the average number of secondary infections that arise from one productively infected person is larger than one (1). This is the basic reproductive number and for Eq. (1) it is defined by *R*_o_=*βσp*/(*δc*). The intrinsic growth rate constant, *r*, is solved for by the dominant root of the equation *r*^2^+(*δ*+*c*)*r*+*δc*(1−*R*_o_). However, if *c*>>*δ* and *r*, then it can be simplified to: *r*=*δ*(*R*_o_−1). When *R*_o_>1, then infection rates will initially experience an exponential increase [41].

The model predicts that as the infection grows it decelerates. The infection will converge in damped oscillations to the non-trivial equilibrium: 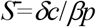, *Ī=*(*R*_o_−1)*δc*/(*βp*), 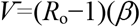. This dynamical steady state is obtained when the number of new infections equals the number of recovering individuals, where every productive infection generates, on average, only one more new secondary infection. A global stability analysis can be found here [42]. As far as we know, this is the simplest dynamical model which affords a non-trivial non-zero infection steady state.

Assuming a quasi-steady state, e.g., the viral dynamics are much more rapid than the epidemiological phenomenon (*p*>>*c*), then Eq. (1) can be reduced to:

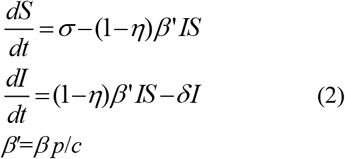

with no loss of generality for the major trajectories of infection dynamics [43]. The model dynamics are shown in Figure 1. This functional form has the advantage to decrease model complexity, especially because the viral compartment is less relevant at the community-scale. Exponential decay under interventions to block infection is given by *r*_0_=*δ*−(1−*η*)*β’S*_0_, where *S*_0_ are the number of susceptibles at *t*_0_. Under highly efficient interventions, i.e., *η*→1, then a minimal estimate for *δ* can be derived directly from the observed decay half-life of t_½_=ln(2)/*δ* [44,45]. When social interventions are withdrawn or vaccines become ineffectual at time *t*_1_, infections rebound at an exponential rate given by *r*=*β’S*_1_−*δ*, where *S*_1_ is the level of available susceptibles at *t*_1_. Crucially, *r* can be obtained directly from the observed slope on the semi-log graph, and its doubling-time is t_2_=ln(2)/*r*. This expansion in infections will continue in damped oscillations returning to the steady-state.

**Figure 1.**
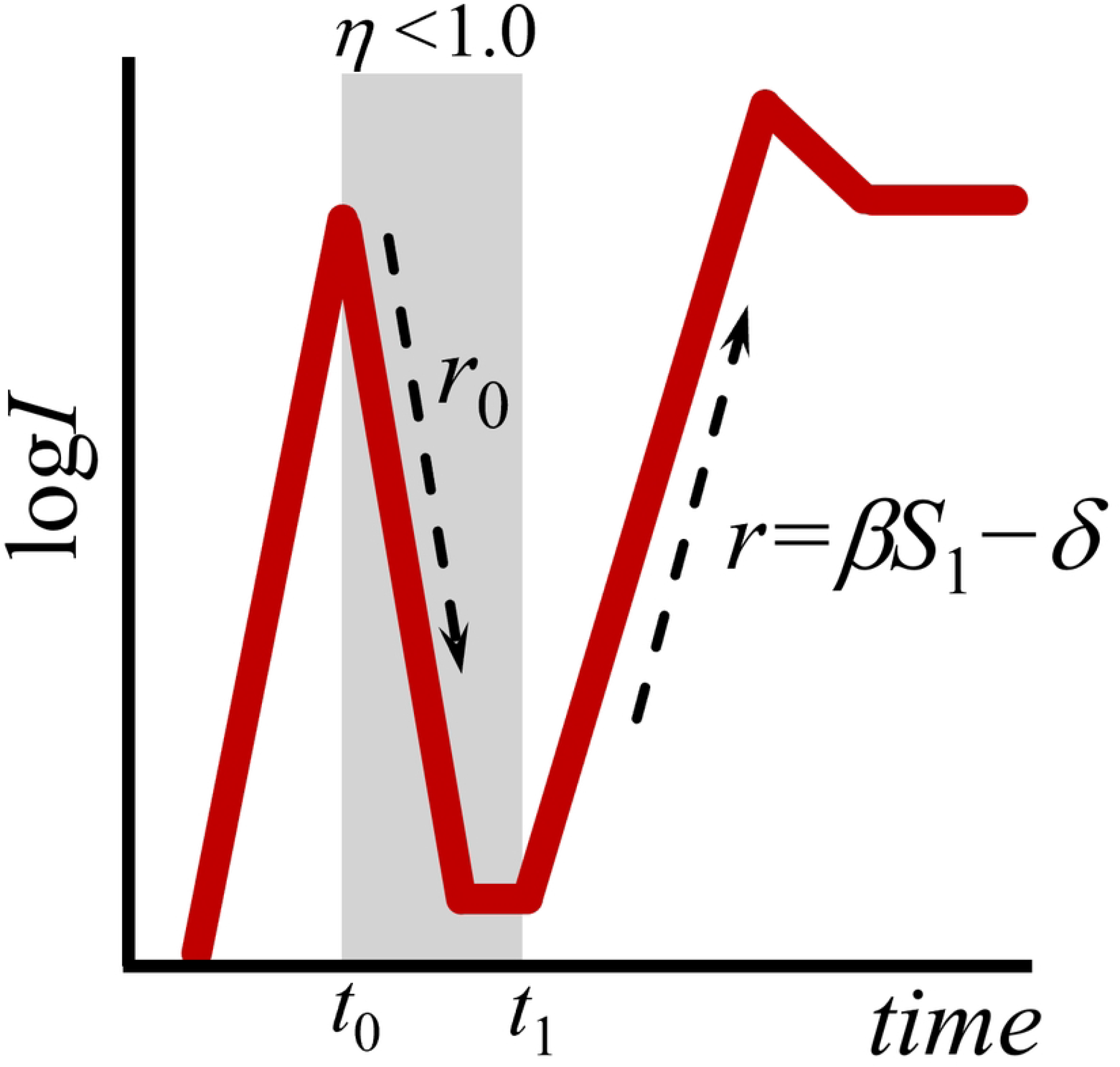
Epidemiological dynamics under interventions to block infection. Initially, infections rise exponentially (though national COVID-19 testing programs were also ramping up). During stringent lockdown and effective cessation of viral transmission, between *t*_0_ and *t*_1_, infection decays exponentially with a half-life of t_½_=ln(2)/*r*_0_, where *r*_0_ is derived from the slope of the ln-transformed infection data. This provides a minimal estimate for the value of parameter *δ*, assuming partial intervention efficacy (0<*η*<1). This decay will decelerate reaching a lower steady state. Infections will naturally rebound upon lifting of interventions and/or loss of vaccine efficacy with a doubling time of t_2_=ln(2)/*r* and *r* also calculated from the exponential up-slope. The system will converge with damped oscillations to an elevated infection steady state. This basic pattern will recur as interventions are deployed at different times.

### Estimation of the basic reproductive ratio

The basic reproductive number is based on a ratio among all five model parameters. However, the paucity of independent knowledge and accurate values for them precludes adequate approximations of *R*_o_. To alleviate this, the relationship between the basic reproductive ratio (*R*_o_) and the exponential growth rate (*r*) can be recovered such that *R*_o_=1+*r*(*r*+*δ*+*c*)/*δc*. If *r*+*δ* is small compared to *c*, then this approaches:

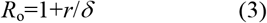

which can be calculated directly from the exponential slopes, *r*_0_ and *r*, as described above.

### Parameter values and statistical analysis

To determine the initial values for model parameters, half-life decay during lockdowns and rebound doubling-times were calculated from the log_n_-transformed data of confirmed cases (weekly geometric means). Optimized values were generated by nonlinear fitting (Berkeley Madonna v8), minimizing the objective function 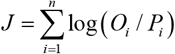 where *O*_*i*_ and *P*_*i*_ are the observed and expected values, for *n* datapoints, with the advantage of stabilizing the variance during the fit [28]. Many functional forms for intervention efficacy (*η*) can be used but for simplicity, generalizability and as a first approximation:

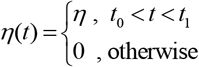

for each intervention wave. The observed decrease in mobility is used here be used as a proxy to estimate its value for each country [46]. Trivially, the proportion of the population needed to be vaccinated in order to block community spread, known as “herd immunity” threshold is [47,48]:

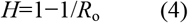

Longitudinal comparisons on the parameter values is performed using the Mann-Whitney u test. 95% confidence intervals, along with their statistical significance, are calculated as appropriate. Model errors (RMS) are reported. Data, simulations and results are available online at: https://github.com/davidville/COVID-19.

## Results

### Dynamics of COVID-19 epidemiology

A preliminary analysis of confirmed COVID-19 cases from 15 nations which did not implement stringent lockdown policies, or were unsuccessful at their implementation, indicates widely varying rates and infection levels (Fig. 2). By the end of Feb 2020 these nations had initial infection levels of ∼10^0^ cases per km^2^ with sustained infection doubling times of 1.2-1.7 weeks. Levels increased exponentially for 20±8 weeks and stabilized around a dynamical steady state with fluctuations no more than 0.5log. Setpoints among these countries were 100- 400 cases per km^2^ built-up area per day. Interestingly, South Africa and Armenia exhibited spontaneously oscillating kinetics with an amplitude one order-of-magnitude, perhaps alluding to the existence of a ‘limit cycle’. India exhibited one of the largest differences in infection over time, increasing to 10^2.5^, declining to 10^1.5^ then peaking at 10^3^ before declining spontaneously again to 10^2^ cases per km^2^ built-up area per day. Because there were no effective measures to block COVID-19 spread, the number of confirmed cases attained a dynamical equilibrium around which case numbers fluctuated.

**Figure 2.**
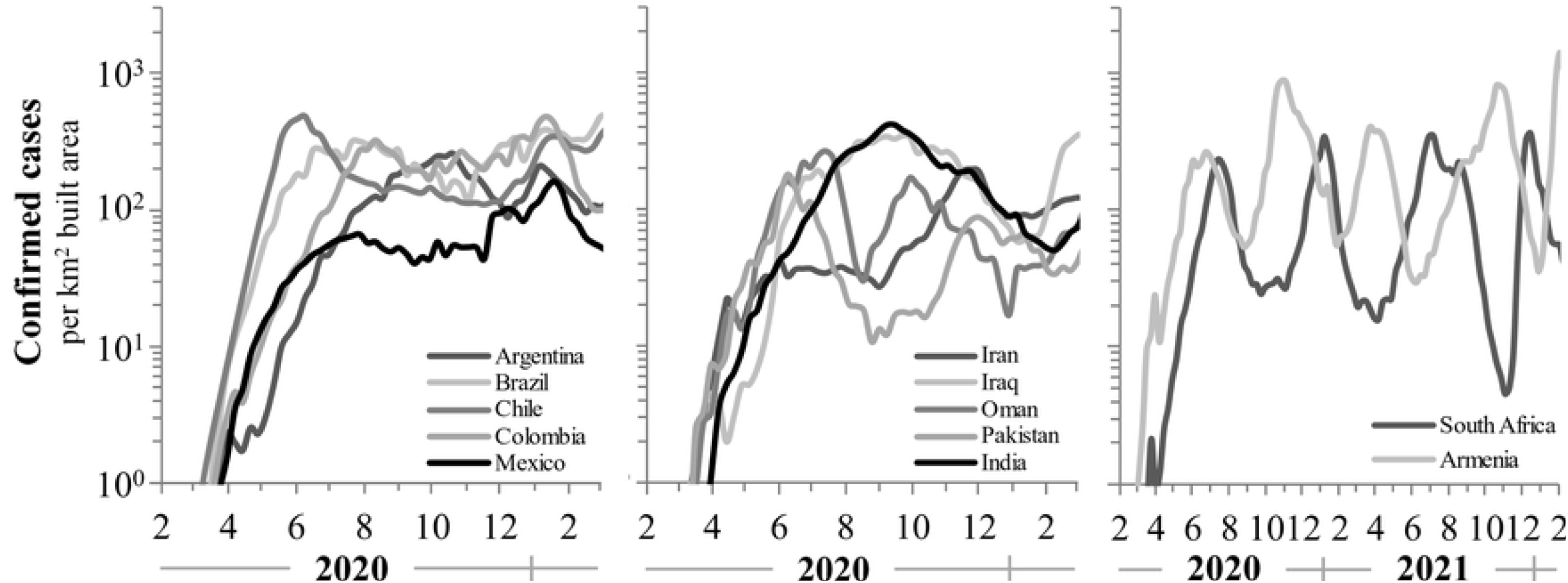
COVID-19 case levels for 10 nations with no or ineffective lockdowns (right and center panels) as well as the Republic of South Africa and the United States (right panel), which instituted effective lockdowns.

**Figure 3.**
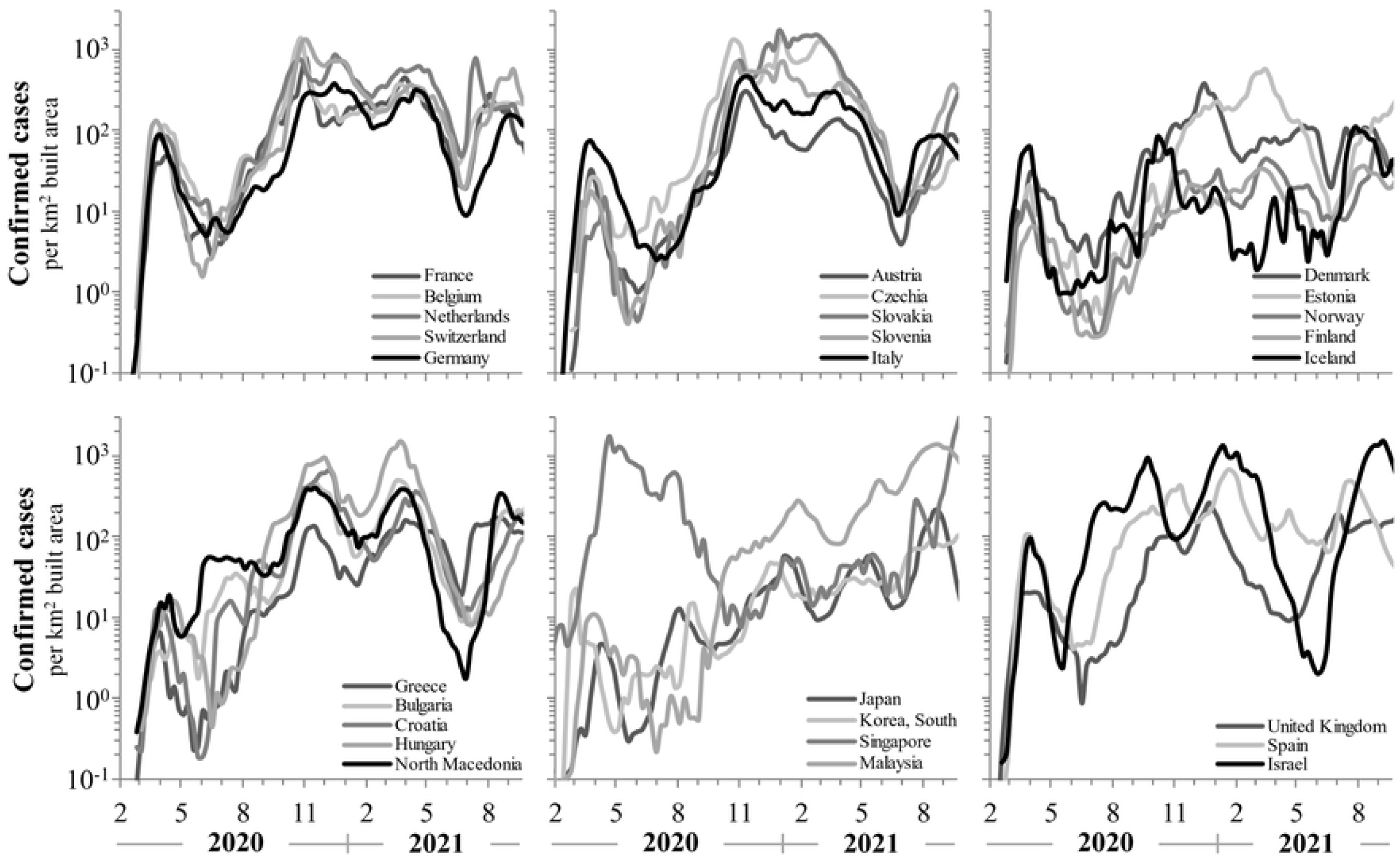
COVID-19 positive confirmed cases between February 2020 and September 2021. Data are normalized to built-up area to account for density effects in infection rates. On this scale the recurring patterns become apparent. The exponential decay during lockdowns and following vaccination is clear, as are the geometric rebound trajectories. On this scale the recurring patterns in COVID-19 community diffusion kinetics are undoubtedly evident.

**Figure 4.**
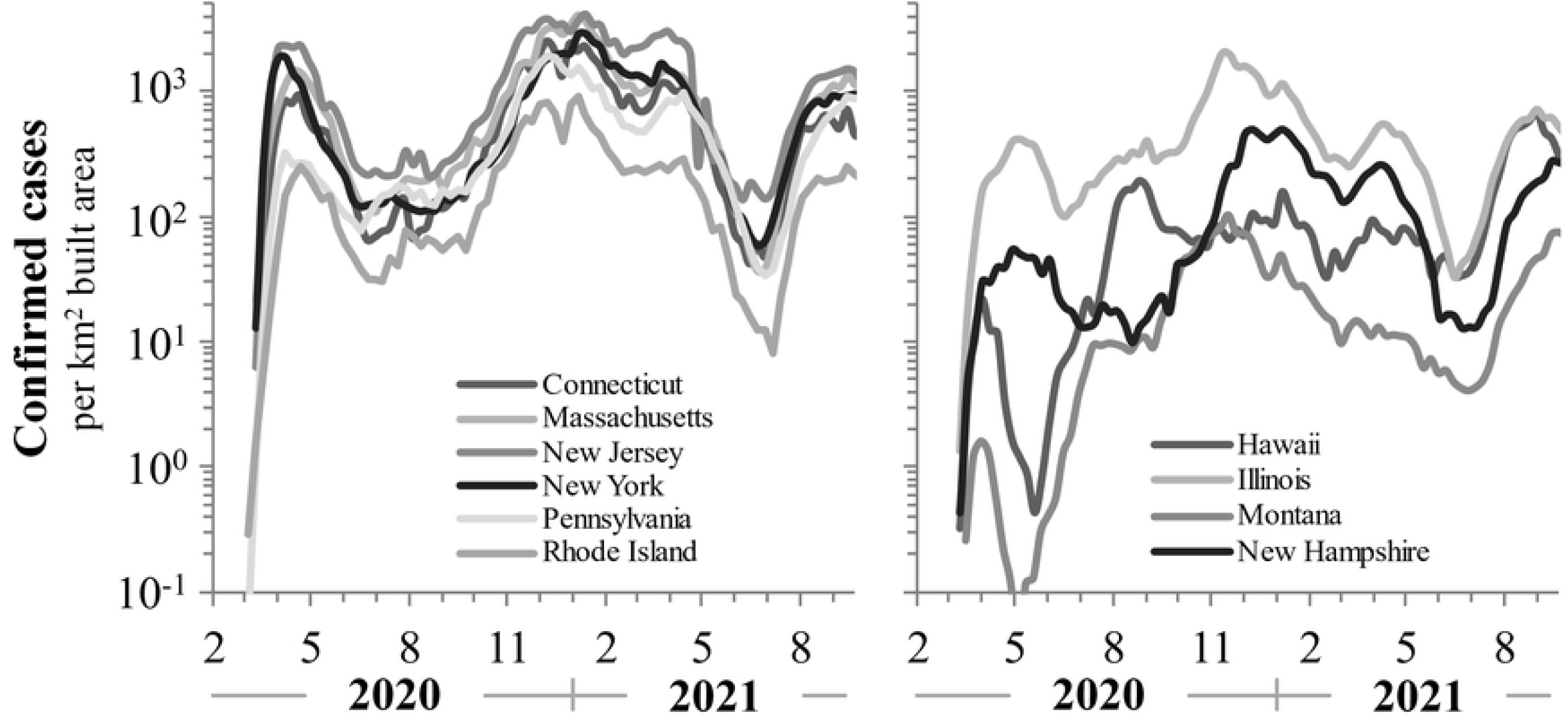
COVID-19 positive confirmed cases ten US states conforming to inclusion criteria from February 2020 to September 2021. More rural and less dense populations have lower COVID-19 infection rates, in general. Data are normalized to built-up area to account for density effects in infection rates.

### Dynamics during effective lockdowns

COVID-19 positive case turnover allows analysis of effective social distancing through population-level lockdowns. Non-pharmaceutical means to block new rounds of infections were initially rapid and effectively implemented. Infections begin to decay exponentially 7-10 days after the lockdown policies are implemented, with down slopes of 0.5±0.3 per week and corresponding to half-life values of 2.0±1.1weeks. Infection rates attained nadir within 4-6 weeks with average efficacy of 68% (range: 46-93%), declining 1-2log lower than pre-lockdown case numbers. Confirmed cases rebounded exponentially with doubling times of 2.3-2.6 weeks following the end of severe lockdowns. The trajectory then converged on an empirical equilibrium steady state of approximately 10^2^-10^3^ cases per km^2^ built area and with fluctuations less than 0.5log.

The USA is composed of distinct political entities, with large inter-state variation. SARS-CoV-2 surged and waned differently, peaking and ebbing at different times among the various states. Therefore, analyses of COVID- 19 for the USA have been done at the state level. Ten states conformed to the inclusion criteria. The US state COVID-19 dynamics were less extreme with lockdown declines of less than 2log in some states, albeit the up- and down slopes during lockdowns were comparable with European nations. Four states suffered elevated steady-states approximately one order-of-magnitude higher (10^3.2^-10^3.5^ cases per built-up area per day).

The UK as a whole had, on average, similar dynamical characteristics as its neighbors. However, the observed decay rates during lockdowns were significantly less rapid, leading to differences that will be expanded upon later. Asian nations, generally, had somewhat different COVID-19 trajectories. While the initial doubling times before lockdowns were similar to other nations and regions, half-lives during lockdowns were nearly twice as rapid, 1.3±0.5 *vs*. 2.0±1.1 weeks. The Asian rebound rates differed less relative to other countries, though they were more prolonged with some clear oscillatory effects. Additionally, the setpoint infection rates in Japan and South Korea were an order-of-magnitude lower than in Europe.

The earliest, most stringent and prolonged restrictions were implemented in Australia and New Zealand (Fig. 5). Confirmed case rates were perturbed to low levels for 35 months. They were kept 0.5log below the lowest rates achieved in Europe until July 2021. Even so, these strict “Zero COVID” policies were insufficient to snuff out community spread entirely. As limits were relaxed, infections surged exponentially with doubling times and equilibrium states comparable to elsewhere, even in the milieu of high vaccination coverage.

**Figure 5.**
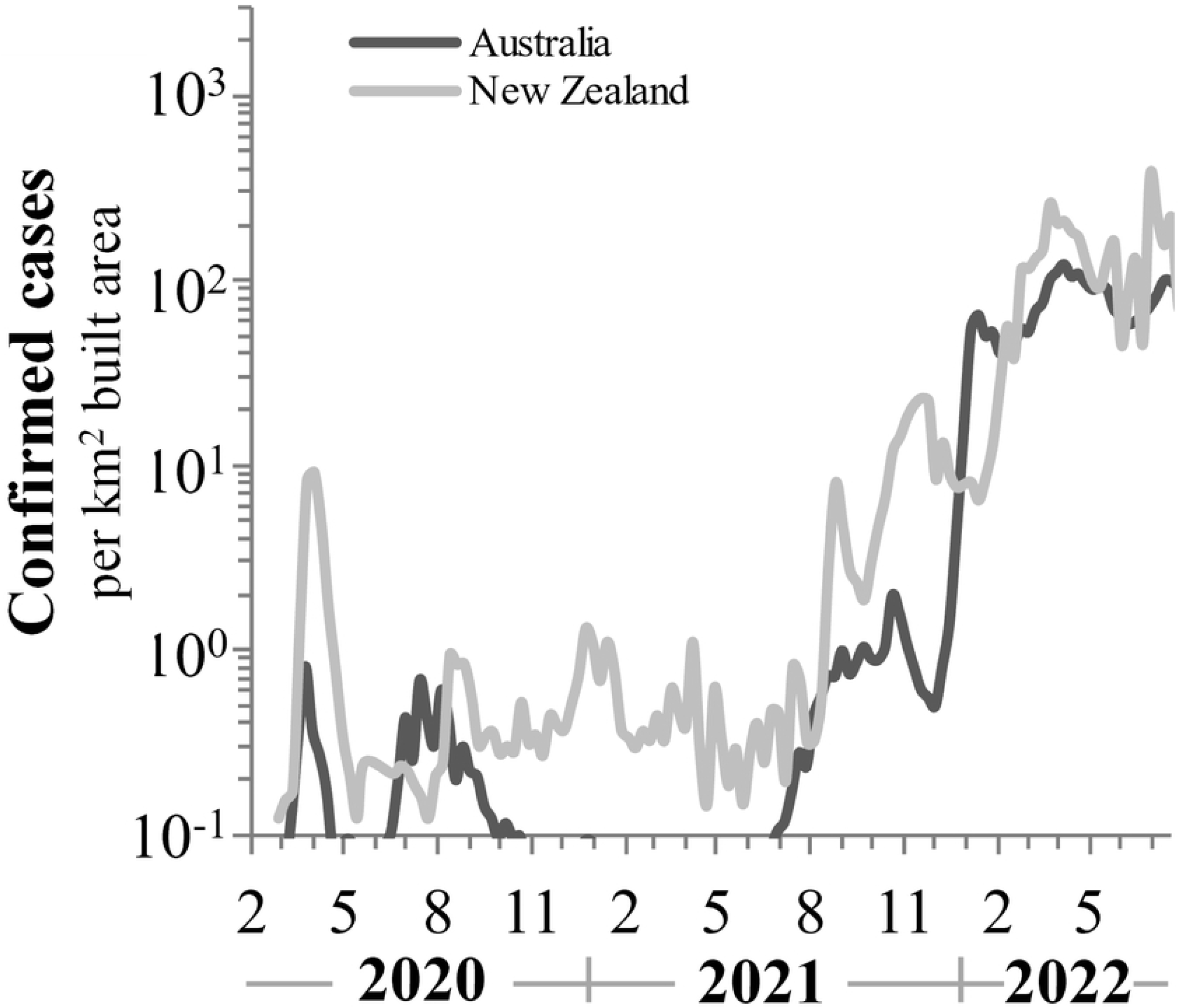
COVID-19 positive confirmed cases for Australia and New Zealand from February 2020 to September 2021. The strict “Zero COVID” policies implemented for 35 months kept infection levels but they rebounded when restriction were lifted and achieved levels similar to those in Europe. Data are normalized to built-up to account for density effects in infection rates.

### Modeling of early COVID-19 infection dynamics

The frequent and robust PCR testing for COVID-19 deployed in nations and regions included here allow for the mathematical analyses of infectious persons. Results of the modeling and the parameter values obtained are found in Table 2. The infection dynamics parameter values were obtained from the exponential slopes directly from the data. Initial infection expansion rate constants were 0.5-0.7 per week during February-March 2020, with corresponding doubling times of 1.2-1.6 weeks. Social distancing and lockdowns resulted in exponential decay of infection rates from the pre-lockdown peak values of 0.4-0.5 per week with half-life values of 1.7-2.3 weeks. This provides a maximal estimate for the case recovery rate constant parameter (*δ*).

Infection rates rebounded with doubling times of 2.6-3.7 weeks (range:0.6-4.4 weeks) upon lifting of the extreme social distancing measures. These represent a minimal estimate for *r*_0_. This is four-fold less rapid than the initial pre-lockdown exponential growth rates. Finally, after 4-12 weeks infections reached a relatively stable setpoint level with values ranging among countries ranging between 10^1.3^-10^3.4^ (CI_95%_: 10^2.3^-10^2.6^) cases per km^2^ built-up area per day. Notably, initial pre-lockdown infection rates are significantly correlated with steady state infection levels (PPMCC=0.41, P=0.037) alluding to the importance of the intrinsic infection rate and extent of very early viral expansion in the infective dynamical and endemic steady state.

Similar patterns were observed for ten states in the USA and five nations in Asian regions. Israel implemented a second lockdown in 2020 leading to infections decaying with a half-life of 1.5 weeks and subsequent rebound with a doubling time of 2.0 weeks; values which are only 15 and 43% more rapid than those during the primary lockdown, respectively. Markedly, not only were decay and rebound slopes among countries of similar magnitude, but they were also similar among infection waves within countries.

### Basic reproductive number (R_o_)

The analytical approach here contributes insight on the basic reproductive ratio for the community spread of SARS-CoV-2. In the literature reporting on COVID-19, and other epidemics, this is approximated from the initial exponential growth phase [14] and represents an overestimation because it ignores the *β/δ* ratio. Here the “natural experiment” of the efficient impedance of viral community spread during the initial phase of the SARS-CoV-2 pandemic allows the use of the empirical rebound up-slope (*r*) and values for the recovery/removal rate constant (*δ*). The estimates for the basic reproductive number are provided in Table 2. Using experimentally established values for *δ* (0.4-0.5) from the decay slope during interventions to block viral expansion and ranges for *r* (0.2- 0.3) leads to basic reproductive numbers ranging between 1.4-2.3, narrowing for a CI_95%_ to 1.6-1.8. From this perspective, active COVID-19 infected individuals would generate approximately 1.7 new secondary infections, on average.

### Herd immunity and inhibition of infection by vaccination

Herd immunity is a threshold value at which new infections cannot perpetuate within the community and is derived from the basic reproductive number. Indeed, nearly all countries experienced a rapid exponential decline in case numbers with efficacies of 44-99% (CI_95%_: 64-72) and half-life values similar to those during lockdowns (CI_95%_: 1.3-1.7. Table A1) following the distribution of SARS-CoV-2 vaccinations. The observed percent of the population vaccinated which coincides with decay in confirmed cases is between 44-55%, based on the nations and regions included here (Table 2). First, empirically, the values for *R*_o_ in other studies seem extremely high. Second, now it is possible to test the previous calculation of *R*_o_, which should be smaller than the observed values. Indeed, the observed “herd immunity” was slightly above the values derived mathematically, as expected from Eq. (4).

### Delta variant wave rebound

In June 2021, after the large decrease in COVID-19 following national vaccination programs, COVID-19 cases rebounded spontaneously. The wave was driven by the Delta variant, which became dominant. This rebound was characterized by doubling times of 1.1-1.3 (Table A1). It finally reasserted levels similar to those observed prior to vaccination deployment. The decay due to vaccinations and this resurgence both correspond to the trajectories observed in 2020.

## Discussion

Infection doubling times (t_2_) and half-life (t_½_) values reveal consistent rates with extremely small variance and narrow range, longitudinally, among all countries analyzed here (Table 1). Mean doubling times for infection levels during the initial exponential phase of the pandemic were 1.0 weeks (CI_95%_: 0.5-2.0). These were quite robust with a caveat about the rate of deployment of testing regimes (see shorturl.at/hmuFN for analysis).

**Table 1.** COVID-19 kinetic characteristics in countries with no effective lockdowns

| Country | Initial growth rate $t_2$ | | Time to steady state weeks | Steady State $\log I \pm \text{SD}$ |
| --- | --- | --- | --- | --- |
| Argentina | 0.2 | 2.8 | 25 | $2.2 \pm 0.3$ |
| Armenia* | 0.4 | 2.0 | 11 | $2.2 \pm 0.4$ |
| Brazil | 0.6 | 1.1 | 13 | $2.4 \pm 0.2$ |
| Chile | 0.4 | 1.6 | 13 | $2.4 \pm 0.3$ |
| Colombia | 0.3 | 2.4 | 21 | $2.4 \pm 0.3$ |
| Costa Rica | 0.5 | 1.3 | 26 | $2.4 \pm 0.5$ |
| Ecuador | 1.3 | 0.5 | 19 | $1.9 \pm 0.1$ |
| El Salvador | 0.4 | 1.7 | 17 | $1.8 \pm 0.3$ |
| India | 1.1 | 0.6 | 20 | $3.9 \pm 0.5$ |
| Iran | 0.2 | 4.1 | 45 | $2.3 \pm 0.4$ |
| Iraq | 0.4 | 1.7 | 22 | $2.5 \pm 0.2$ |
| Mexico | 0.8 | 0.8 | 17 | $1.7 \pm 0.2$ |
| Oman | 0.5 | 1.5 | 11 | $2.1 \pm 0.4$ |
| Pakistan | 0.3 | 2.6 | 25 | $2.3 \pm 0.4$ |
| Peru | 0.8 | 0.8 | 16 | $2.5 \pm 0.3$ |
| S. Africa* | 0.4 | 1.9 | 16 | $1.8 \pm 0.4$ |
| **mean** | **0.5** | **1.7** | **20** | **2.3** |
| **CI<sub>95%</sub>** | **0.4-0.7** | **1.2-2.2** | **15-24** | **2.0-2.6** |

\*) limit cycle dynamics

**Table 2.**
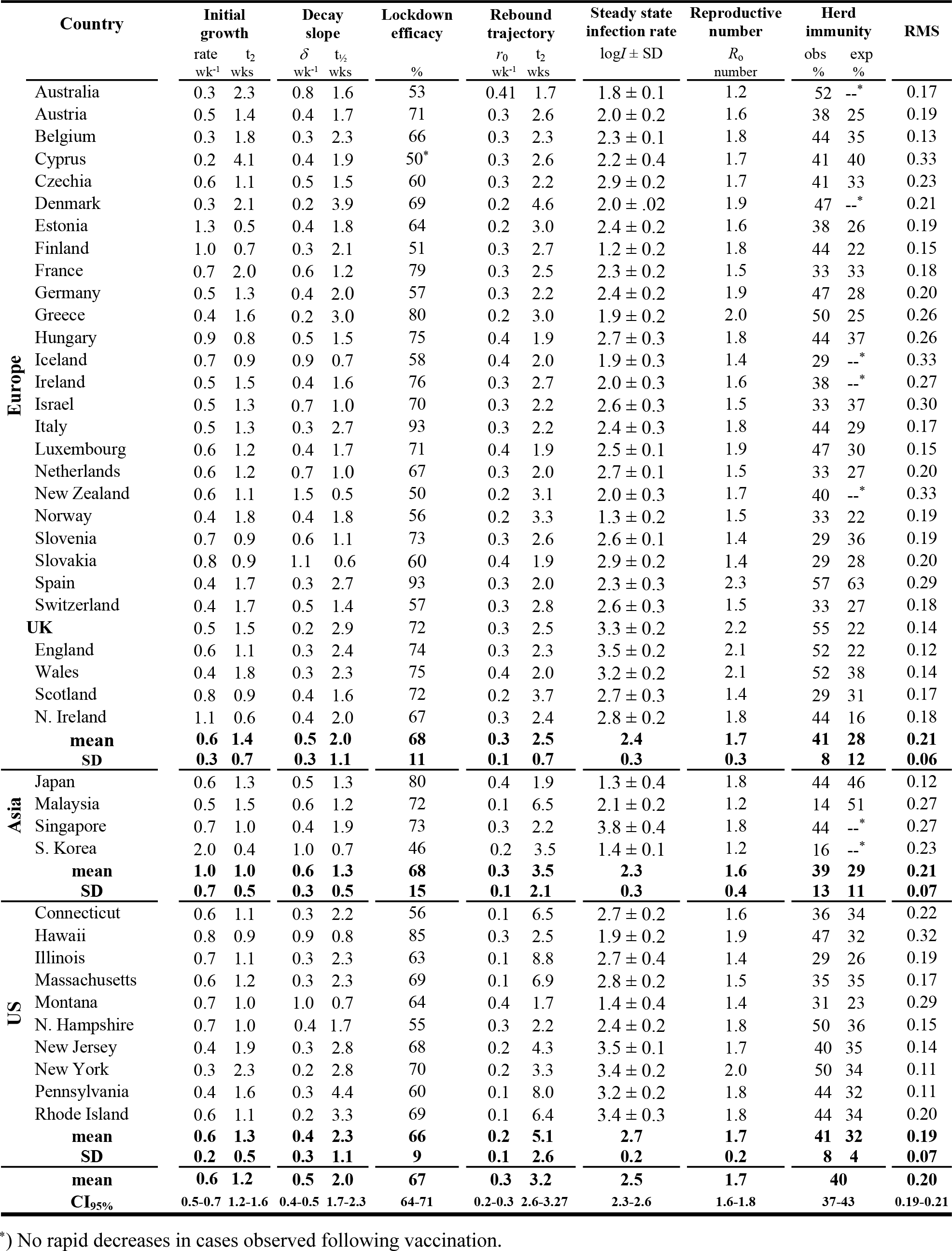
COVID-19 model characteristics

Lockdowns were extremely effective by inhibiting physical contact and blocking the virus from circulating. Countries with no effective social distancing measures rapidly reached a setpoint equilibrium state. Limiting movement of the population was related to lockdown efficacy. Restrictions to travel of 45-93% decreased infection rates by 10-fold or more, leading to an exponential decay of >90% in confirmed cases. Importantly, this was uncorrelated with the minimal infection numbers. More stringent lockdowns do not appear to confer further inhibition to stop viral diffusion and may signify the existence of an optimum in interventions to block COVID- 19. The mean associated half-life value during lockdowns was 2.0 (CI_95%_: 1.7-2.39) with no statistically significant difference among the nations and regions studied here. The epidemiological interpretation of this measure is the maximal value for the recovery rate of infected individuals.

As distancing policies were lifted, infections rebounded exponentially as viral diffusion over the social network is no longer perturbed. Intrinsic doubling times can therefore be determined empirically by the up-slope on a semi-log graph. The observed doubling time was consistent with 2.5±0.7 weeks in European countries. Asian nations included here had values of 3.5±2.1, perhaps owing to their stricter regulations. In the states of the United States the value was even higher at 5.1±2.6 weeks.

Taken together, the rebound and decay rates were harnessed to provide a maximal estimate for the basic reproductive number. *R*_o_ is consistent with a mean value of 1.7 (CI_95%_: 1-6-1.8), due to the invariance of the model parameters. Spain, Greece, and Britain (*i*.*e*., England and Wales) were areas with elevated infectivity with values of 2.3, 2.0 and 2.1, respectively. An important outcome of this calculation is the elucidation of the epidemiological “herd immunity” threshold.

During emergent pandemics, estimates of the basic reproductive number tend to be overestimated. EarlyCOVID- 19 studies reported very high values [49,50]. Our estimates for SARS-CoV-2’s *R*_o_ vary only slightly during waves of COVID-19, which would make sense if the dynamical properties of the infection did not appreciably change, and they are comparable to historical influenza pandemics [51] and commensurate with seasonal influenza outbreaks [52]. Although these estimates are substantially lower than those reported elsewhere for COVID-19, they agree with some studies [53].

Vaccination deployment against SARS-CoV-2 had a dramatic effect on infection rates. Confirmed cases decayed exponentially with a mean half-life value with similar rates as during the social distancing lockdowns, after achieving the herd immunity threshold. For example, Israel with its early and rapid program experienced a half-life of 1.03 weeks in confirmed cases once 45% of the population was immunized. This agrees with the prediction given by the approximations for *R*_0_ based on Eq. (3).

Following the achievement of herd immunity, after approximately 30 weeks, infections spontaneously rebounded again as the delta-variant emerged. The observed escape trajectory was empirically equivalent to the rebound trajectories following the lockdowns and with doubling times approximately every 1.2±0.3 weeks, similar to the post-lockdown rebounds doubling times. Interestingly, the Delta variant emerged in every nation included here within 4 weeks, surprising due to the low volume of international travel. Finally, infection rates returned to similar levels as the pre-vaccination setpoint and invariant among the sampled countries.

Although infection rates tended to initially increase exponentially when numbers were low, they quickly saturated to a level of 10^2^-10^3^ confirmed cases per km^2^ built-up area per day. This was reached in nearly all nations and regions within 4-6 weeks, even in absence of interventions. Even New Zealand and Australia with strict and highly effective lockdowns rapidly reached this level of infections with the lifting of social distancing measures). Such observations, seen everywhere, suggest a basic, perhaps fundamental, shared epidemiological dynamic and the importance of population density for the spread of SARS-CoV-2 [38,54,55].

As we have shown, waves of both infection and suppression can define COVID-19. Our concluding perspective views the infection data decomposed into their wavelet phases and modeled with the generalized multi-logistic model [56]. This approach allows derivation of the saturation level of cases as well as the “characteristic time” (*Δt*) denoting how long the infection takes to increase from 10% to 90% of its extent. While data for many nations and regions resolve neatly into a succession of waves, Israel is unusual in having excellent data for (so far) seven waves of infection as well as companion data about societal responses and suppression for the first five waves. Figure 6 shows the first five infection waves and their durations ranging from 4.4 to 10.6 weeks. The sequence of waves suggests the extremely dynamic interaction of COVID-19, generating new variants, with the social and medical context, including lockdowns, distancing, and vaccines. Predicting new waves remains an unsolved challenge.

**Figure 6.**
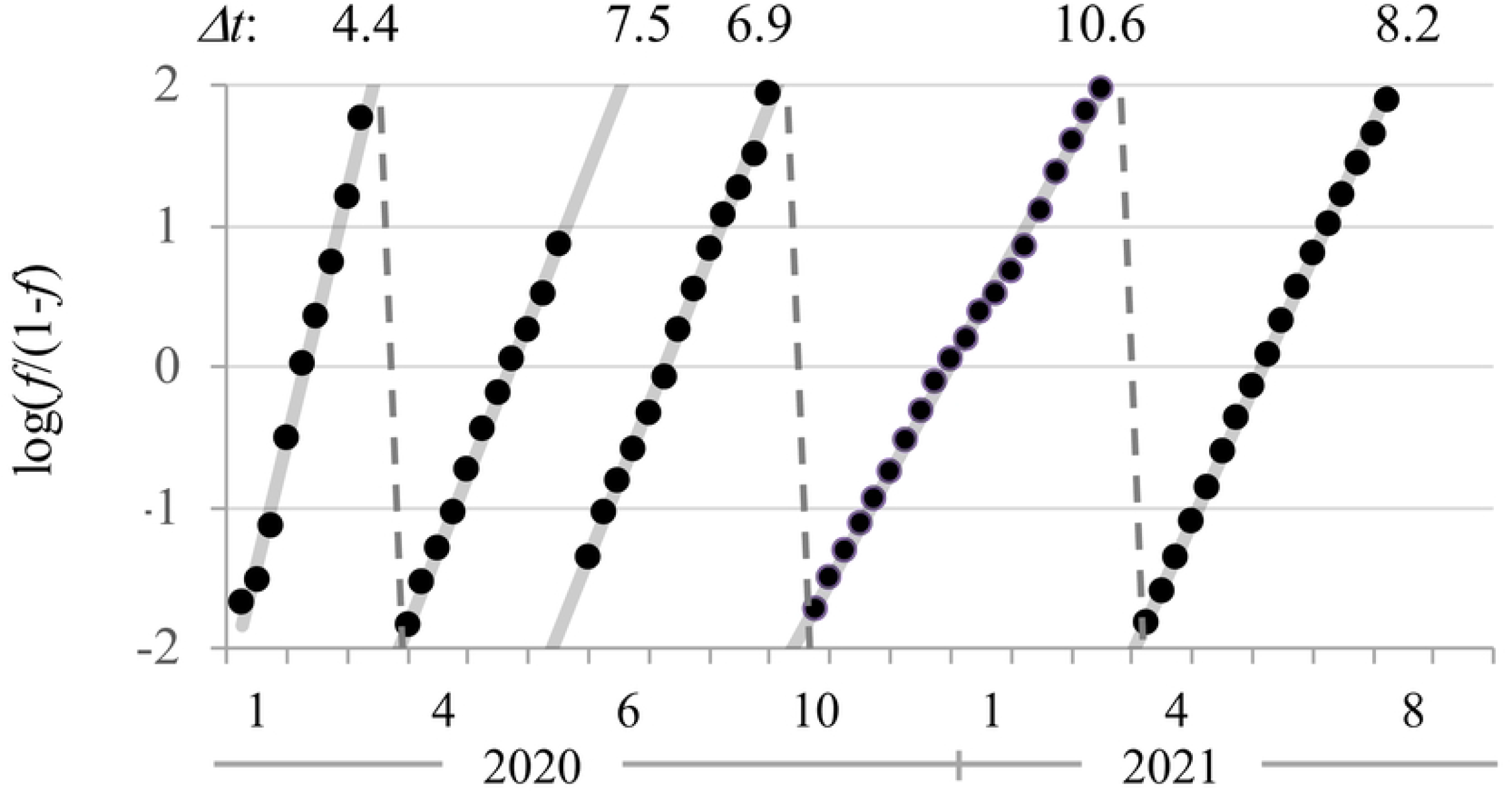
Logistic curves for first five waves of COVID-19 in Israel and the number of weeks each waves took to run its course.

To conclude, the dynamical properties of COVID-19 epidemiology are conserved with consistent kinetic patterns with little variation during multiple waves of infection and globally among nations and subnational regions. Nations and regions which implemented lockdowns sufficient to block community spread effectively experienced a rapid decline in confirmed cases. However, with lifting of interventions, rates rebounded to the previous high infection rates and attained a relatively stable empirical steady state. For COVID-19 societies so far appear to face a choice between relatively high oscillations involving waves of suppression and infection and lesser oscillations around an endemic setpoint. The approach presented here based on the viral dynamics paradigm allows derivation of fundamental measures vital to policy such as the basic reproductive number and lockdown efficacies. Values for *R*_o_ derived here of 1.6-1.8 are maximal estimates and lower than other reports. Information on variables of interest for policy normally difficult to obtain is available through this approach and may suggest monitoring strategies efficient for accurate determination of the dynamical properties of future pandemics.

## Data Availability

https://github.com/davidville/COVID-19

## Acknowledgments

We thank Dr. Yoav Dvir for his helpful suggestions. Dr. Mark Y. Stoekle and Dr. David S. Thaler gave important discussions and Ms. Michele Filgate was indispensable on editing the final manuscript.

## Appendix A

**Table A1.**
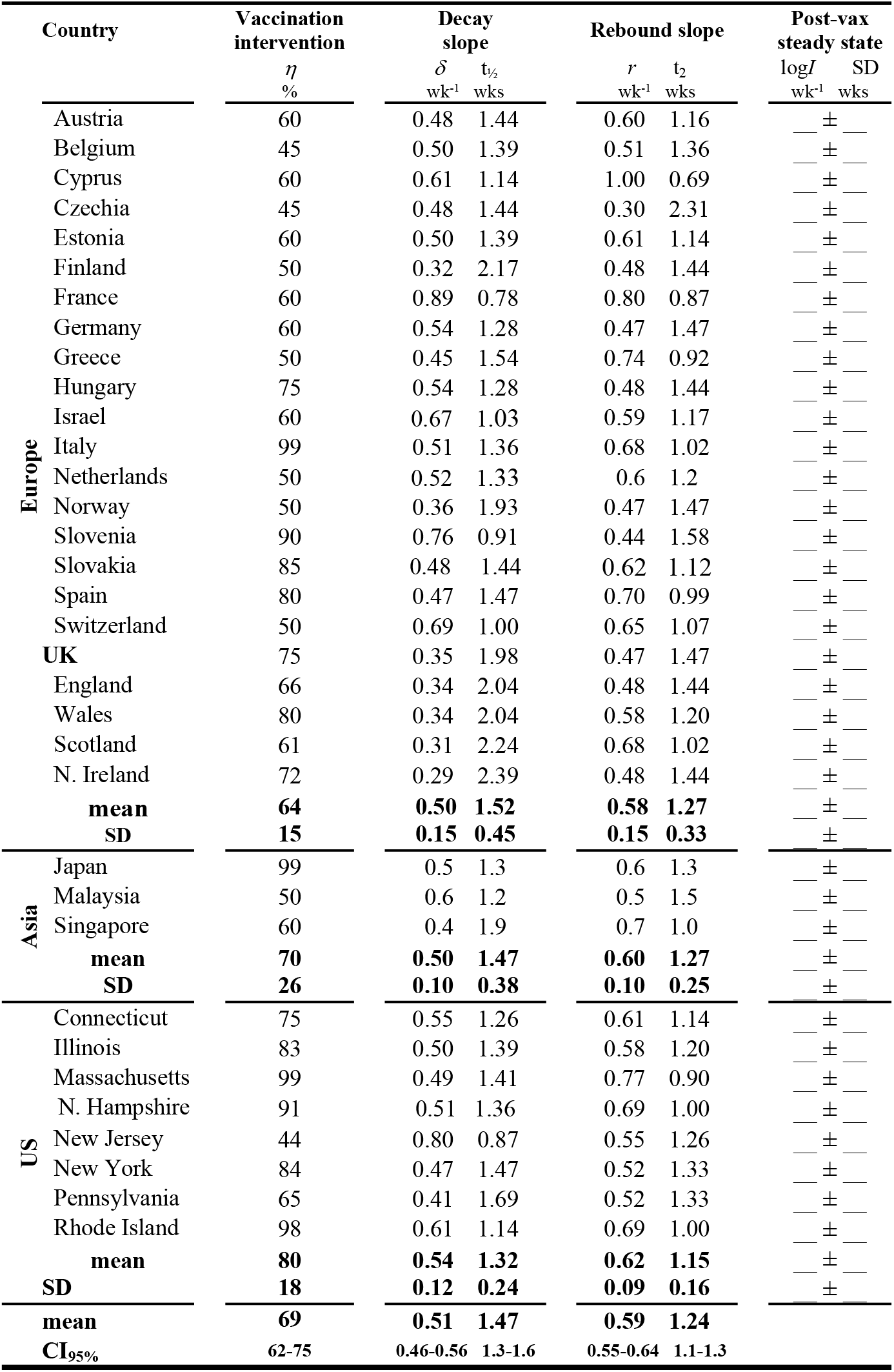
Vaccination decay and Delta variant escape trajectories.

